# Framework for Identifying Drug Repurposing Candidates from Observational Healthcare Data

**DOI:** 10.1101/2020.01.28.20018366

**Authors:** Michal Ozery-Flato, Yaara Goldschmidt, Oded Shaham, Sivan Ravid, Chen Yanover

## Abstract

**Objective:** Observational medical databases, such as electronic health records and insurance claims, track the healthcare trajectory of millions of individuals. These databases provide real-world longitudinal information on large cohorts of patients and their medication prescription history. We present an easy-to-customize framework that systematically analyzes such databases to identify new indications for on-market prescription drugs.

**Materials and Methods:** Our framework provides an interface for defining study design parameters and extracting patient cohorts, disease-related outcomes, and potential confounders in two observational databases, covering more than 150 million patients. It then applies causal inference methodology to emulate hundreds of randomized controlled trials (RCTs) for prescribed drugs, while adjusting for confounding and selection biases. After correcting for multiple testing, it outputs the estimated effects and their statistical significance in each database.

**Results:** We demonstrate the utility of the framework in a case study of Parkinson’s disease (PD) and evaluate the effect of 259 drugs on various PD progression measures. The results of these emulated trials reveal a remarkable agreement between the two observational medical databases for the most promising candidates.

**Discussion:** Estimating drug effects from observational data is challenging due to data biases and noise. To tackle this challenge, we integrate causal inference methodology with domain knowledge and compare the estimated effects in two separate databases.

**Conclusion:** Our framework enables a systematic search for drug repurposing candidates by emulating RCTs that use observational data. The high level of agreement in the results obtained in two separate databases provides an internal validation of identified effects.

## Background and Significance

Drug repurposing, or repositioning [1], is the quest to identify new uses for existing drugs. It holds great promise for both patients and industry, as it significantly reduces the costs and time-to-market of new medications compared to *de novo* drug discovery [2]. To date, the most notable repurposed drugs have been discovered either through serendipity, based on specific pharmacological insights, or using experimental screening platforms [2,3]. To accelerate and increase the scale of such discoveries, numerous computational methods have been suggested to aid in drug repurposing (see reviews in [3–8]). For example, a popular approach, which can be applied to different data types, represents drugs and/or diseases as feature vectors (aka “signatures” or “profiles”), and measures the similarity between these entities or trains a prediction model for drug-disease associations.

In the healthcare domain, the term “real-world data” refers to information collected outside the clinical research settings; for example, in electronic health records (EHRs) or claims and billing data [9]. Such data offer important advantages in terms of volume and timeline span, alongside some inherent challenges such as data irregularity and incompleteness. Recently, real-world data have been increasingly leveraged for various healthcare applications [10]. In the context of drug repurposing, observational data are increasingly used to provide external validation to existing drug repurposing hypotheses. For example, Xu *et al*. [11] used EHR data to validate the association of metformin with reduced cancer mortality. In contrast, there are far fewer examples for utilizing observational data to *generate* new drug repurposing hypotheses. Paik *et al*. [12] derived drug and disease similarities from EHR data and then combined these similarities to score drug-disease pairs and suggest novel drug repurposing hypotheses. Kuang *et al*. [13] leveraged patient-level longitudinal information available in EHRs and applied the Self-Controlled Case Series study design, widely used to identify adverse drug reactions [14], to identify new drugs that can control fasting blood glucose levels.

Conducting a randomized controlled trial (RCT) to validate the efficacy of a candidate drug is costly and lengthy. We propose a framework for generating repurposing candidates by *emulating* RCTs for on-market drugs using observational real-world data. We apply causal inference methodologies to correct for confounding bias, non-compliance to prescribed treatment, and informative censoring [15]. Our framework is configurable, allowing for the specification of inclusion criteria, disease outcome, and potential confounders.

As a test case, we applied the described drug repurposing framework to Parkinson’s disease and emulated RCTs for hundreds of drugs, estimating their effect on three disease progression outcomes. To obtain robust results, we evaluated the agreement of the estimated effects across different causal inference methods and databases. We focus here on general methodological aspects of our framework and the means to validate results. A discussion of the identified drug candidates for Parkinson’s disease and their clinical validity is beyond the scope of this paper. To the best of our knowledge, our study is the first to demonstrate systematic screening for drug repurposing candidates in observational data using RCT emulation, and to compare drug repurposing hypotheses generated in different data resources.

## Materials and Methods

Each emulated RCT estimates the efficacy of a single drug by comparing patient outcomes in two cohorts: drug-treated patients (the “treatment cohort”) versus controls; and correcting for biases between these cohorts, as well as biases related to treatment compliance and incomplete follow-up. In the following sections we describe the study design of the target trials, which largely follows the protocol in [15], as well as our emulation framework.

### Data Sources

We analyzed two individual-level, de-identified medical databases. The IBM Explorys Therapeutic Dataset (hereinafter referred to as Explorys) included the medical data of over 60 million patients, pooled from multiple different healthcare systems, primarily clinical electronic heath records (EHRs). The IBM MarketScan Research Databases (hereinafter referred to as MarketScan) contained healthcare claims information for the years 2011 to 2015. We note that 5.5 million patients (<10%) were known to be covered by both Explorys and MarketScan. As their data originated from different providers, we consider them two separate resources and assume that the overlap in the derived patient cohorts and timelines is negligible.

### Study Design

#### Key Dates

The beginning of the treatment, or its alternative in the trial, is termed the *index date*, corresponding in our emulated RCTs to the first prescription date of the assigned drug. We refer to the observed time before the index date as the *baseline period* and use the information collected during that time period to determine whether a patient is eligible for the target trial. The time following the index date is termed the *follow-up period*, during which the effect of the drug is evaluated. In the Parkinson’s disease (PD) case study, we set the follow-up period to two years. For each patient, we defined *end-of-treatment-compliance* as the end-date of the last prescription of the drug during the follow-up period. We considered patients as censored at the end-of-treatment-compliance. When the prescription duration was unavailable, we set it to three months, the modal value for prescription length in our data. Finally, in the PD case study, we defined *PD initiation date* as the first PD diagnosis or the earliest levodopa (a drug typical to PD) prescription within the year preceding the first PD diagnosis. Since PD is a neurodegenerative disorder that evolves over many years, we allowed the PD initiation date to be moved up by six months. Technically, an earlier (presumable) PD initiation date increased the cohort size in our emulated RCTs, since we required the PD initiation date to precede the index date (see section Eligibility Criteria). Figure 1 illustrates the key dates in our study design.

**Figure 1.**
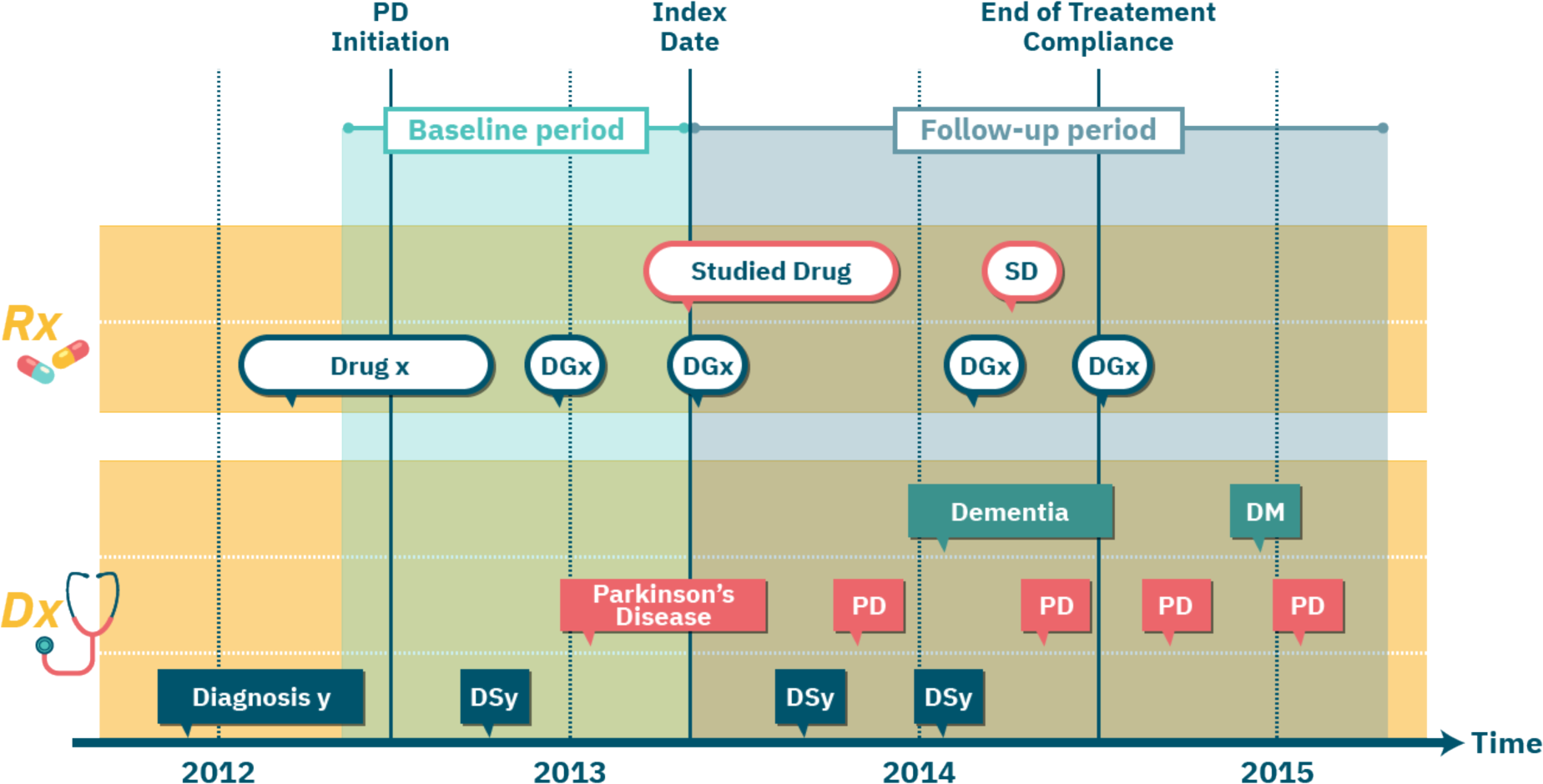
An illustration of the per-patient key dates in the study design of emulated RCTs. Each row corresponds to a certain type of medical event. Ovals indicate diagnosis (Dx) events; rectangles indicate prescription (Rx) events; event type is specified in the first (i.e., leftmost) event in each row and then abbreviated (e.g., “SD” in top row is the abbreviation for “Studied Drug”).

#### Eligibility Criteria

The target trials that we emulated focused on patients suffering from late-onset PD since early-onset patients present different clinical profiles [16,17]. We identified the late-onset patients in our data based on diagnostic codes (International Classification of Diseases, ICD, codes ninth and tenth revision) and required at least two PD diagnoses on distinct dates. Patients diagnosed with PD before the age of 55 were excluded from our study. To allow proper characterization of the patients in our trials, we further required an observed baseline period of one year. To ensure that all “recruited” patients have PD, we demanded that the PD initiation date precede the index date.

#### Treatment Assignment

For a given trial drug, the framework provides two possible settings for defining the control cohort. In the first setting, we use the Anatomical Therapeutic Chemical (ATC) classification system and set the alternative treatment to drugs from an ATC class of the trial drug, excluding the drug itself. In the second setting, the alternative treatment is a drug randomly selected for each patient from his/her list of prescribed drugs; additional criteria can be applied to limit this random drug set. See Discussion for the rationale behind these settings.

For both treatment and control cohorts, we demanded that the assigned treatment had at least two prescriptions ^3^ 30 days apart. Finally, the framework excludes from the control cohort any patient with a prescription for the trial drug. The choice of control is configurable. In the PD case study demonstrated here, we tested both ATC-based and random-drug controls described above. In the ATC-based control, we used the second-level ATC class of each drug, noted as ATC-L2, which includes drugs of the same therapeutic indication. In the random-drug control, we considered drugs that are *not* indicated for PD.

### Outcomes

The efficacy of a drug during the follow-up period is measured with respect to a patient-specific disease outcome, such as the occurrence of a disease-related event. In the PD case study, we defined a set of clinically-relevant events linked to the progression of PD along different axes:

- Fall: as a proxy to advanced motor impairment and dyskinesia.
- Psychosis: measuring progression along the behavioral axis.
- Dementia onset: measuring progression along the cognitive axis (and excluding patients with prior dementia diagnosis from the trial).

We used ICD codes to detect these events (see [18] for details).

### Hypothesized Confounders

Confounders are variables affecting both the assigned treatment and the measured outcome, thus creating a “backdoor path” [19] that may conceal the true effect of the drug on the outcome. Causal effect estimation attempts to block these backdoor paths by correcting for confounders. Since, by definition, confounders influence the treatment assignment, they are computed over the baseline period. In the PD case study, our list of hypothesized confounders contained demographic factors (age and gender), and variables indicating past diagnoses, prescribed drugs, healthcare services utilization, and insurance types.

### Framework for RCT emulation

Our configurable framework follows the study design protocol described above and automatically emulates a maximal number of RCTs using observational healthcare data. Figure 2 shows an overview of the framework and its RCT emulation pipeline.

**Figure 2.**
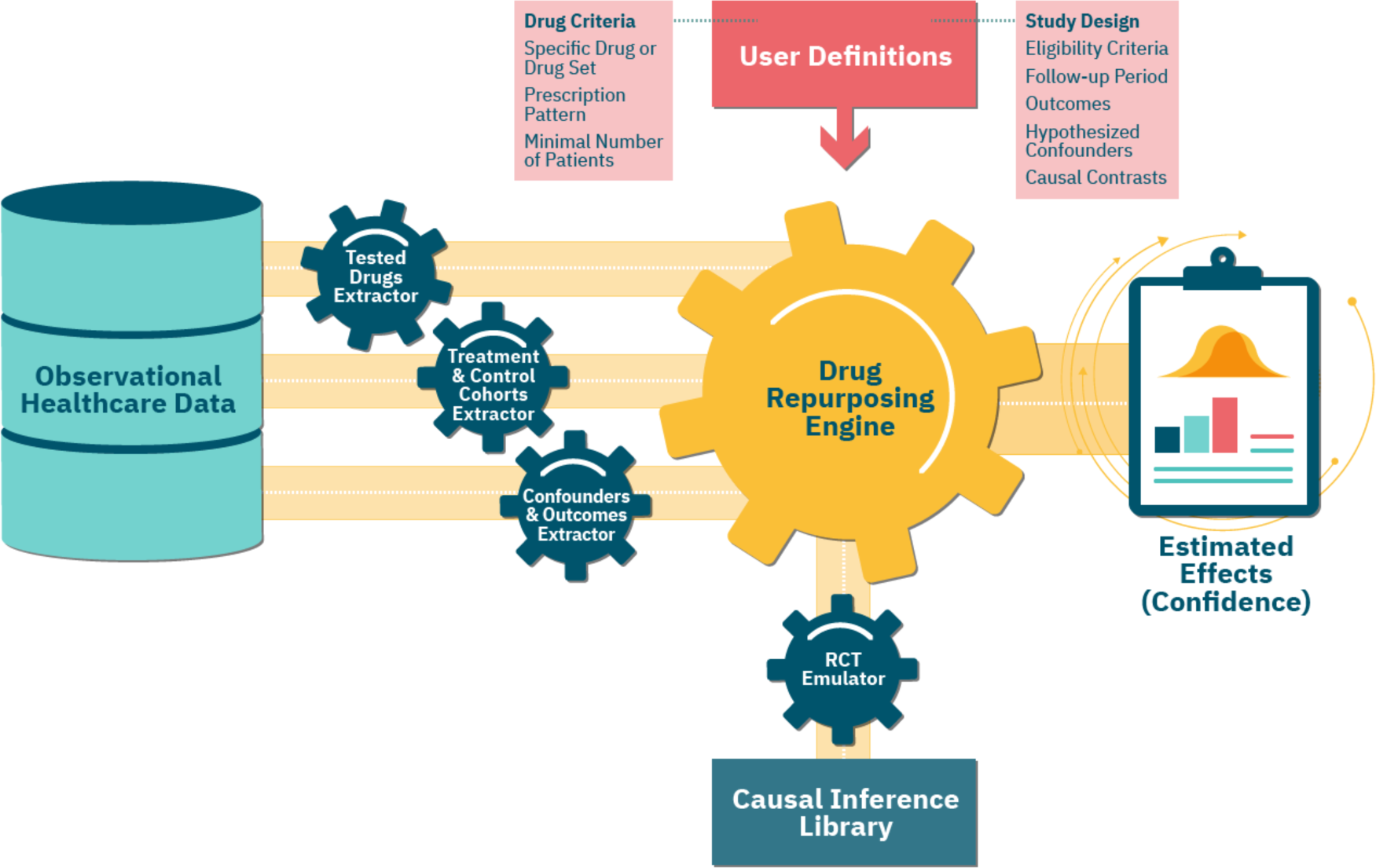
An overview of our framework’s emulation pipeline and the underlying components. First, the *Tested Drugs Extractor* identifies a list of repurposing candidates, based on the user-provided *Drug Criteria*. For each such candidate, using the input *Study Design* parameters, the *Treatment & Control Cohorts Extractor* assigns patients to the respective cohorts. The *Confounders & Outcomes Extractor* computes a baseline and follow-up attributes for patients in both cohorts. The *Drug Repurposing Engine* then instantiates an *RCT Emulator* for each candidate, which estimates its effect on disease outcomes in the treatment versus control cohorts, adjusting for the extracted confounders and using methods implemented in the *Causal Inference Library*.

### Extracting Tested Drugs

Our framework tests all drug ingredients that satisfy the following conditions: (a) it is an active ingredient; (b) it is not part of over-the-counter medications, which may have limited coverage in our data; and (c) the number of patients in the corresponding treatment cohort is above a specified minimal value. In the PD case study, the minimum cohort size was 100 patients. The Tested Drugs Extractor module identifies all the drugs that meet these requirements (Figure 2).

### Extracting Treatment and Control Cohorts

For each emulated trial, our framework uses the feature extraction tool described in [20] to extract the corresponding treatment and control cohorts, and to formulate and compute the values of the confounders and outcomes. In the random-drug control setting, the randomization process is shared by all trials, leading to a large overlap between the control cohorts, and allowing a joint extraction of the confounders and outcomes in these cohorts.

### Emulating an RCT

Below we provide a mathematical formulation of the estimated effects and elaborate on the steps our framework takes to evaluate them.

Let *P*^trial drug^(outcome) denote the expected prevalence of patients experiencing an outcome event in an extreme scenario where *all* patients in the trial (i.e., treatment and control cohorts) were assigned and fully adhered to the trial drug during a *complete* follow-up period. Similarly, let *P*^alternative treatment^(outcome) denote the expected prevalence of the outcome for the analogous extreme scenario corresponding to the alternative treatment. Note that the evaluation of *P*^trial drug^(outcome) and *P*^alternative treatment^(outcome) may greatly deviate from *P*(outcome|treatment cohort) and *P*(outcome|control cohort), namely, the observed (uncorrected) outcome prevalence in the treatment and control cohorts, due to biases in treatment assignment, non-compliance, and loss-to-follow-up. The effect of the trial drug on the outcome is then measured by the difference^1^

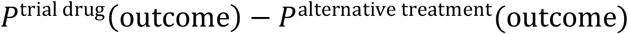

Procedure 1 estimates the effect of the trial drug on an outcome, as well its statistical significance. The steps in this procedure are implemented in the Causal Inference Library module. Details are provided in the following section.

#### Procedure 1: RCT Emulation

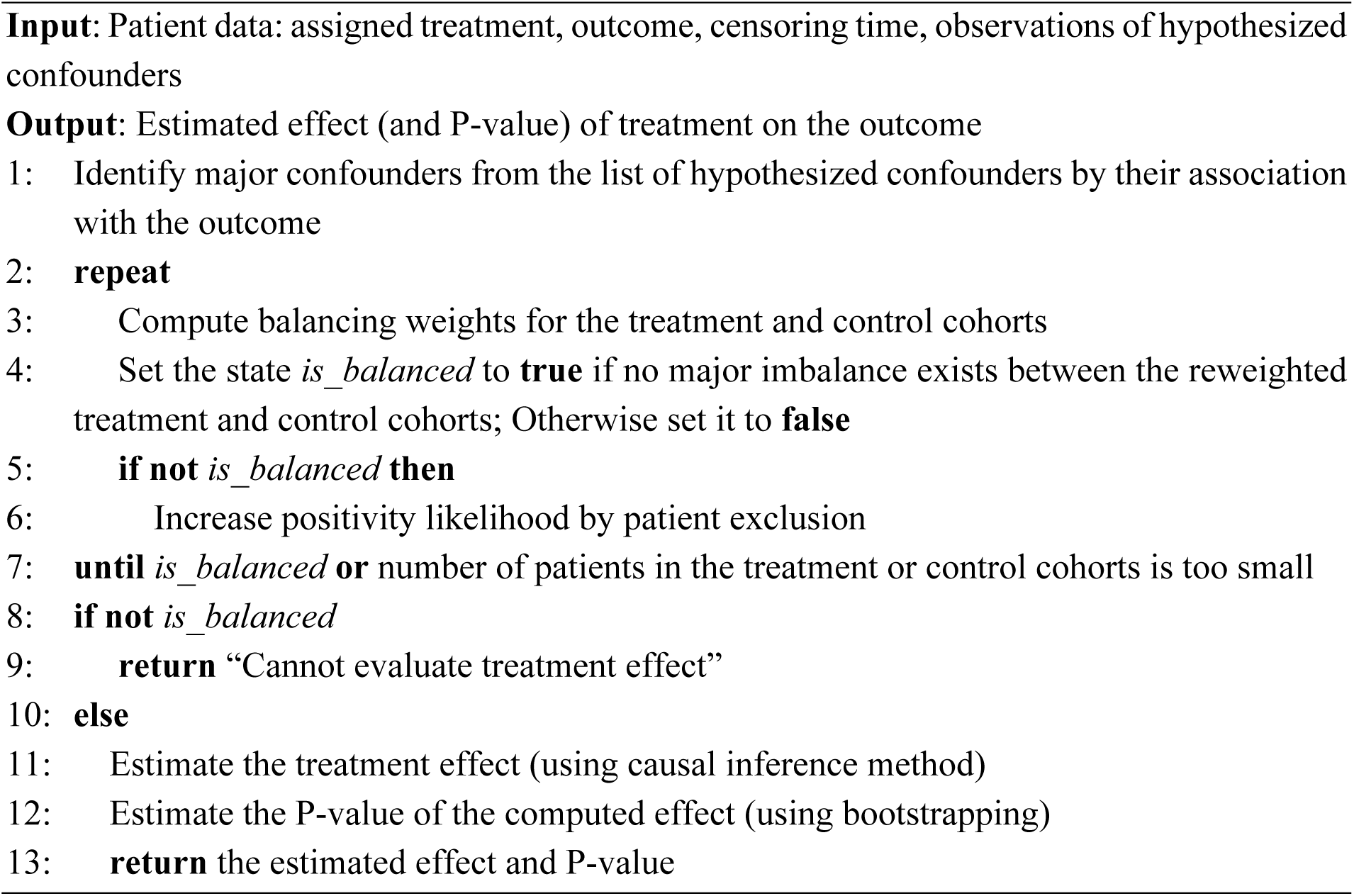

### Causal Inference Library

This module contains various methods to estimate the expected outcomes and causal effects from observational data. For event-based outcomes, as in the PD case study, it offers causal survival analysis methods that adjust for both confounding and selection bias due to incomplete treatment period. Below we provide a summary of the methods used in the PD case study.

#### Identifying Major Confounders

This step identifies major confounders within the set of extracted potential confounders by testing their association with the outcome [21] (Step 1; see also Discussion). First, it excludes features that are nearly constant (mode frequency > 0.99) in both the treatment and control cohorts. It then dichotomizes non-binary feature values into high/low using their median, and measures the association between the feature and the outcome using the following difference

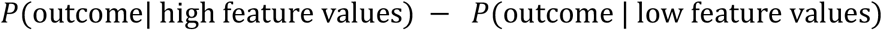

For event-based outcomes, we computed this difference with Kaplan-Meier estimators and used bootstrapping to assess its statistical significance [22]. In the PD case study, we used a P-value ≤ 0.005 in all emulated trials to identify major confounders.

#### Generating Balancing Weights

To generate balancing weights (Step 3), we applied the popular method of inverse probability weighting (IPW) with stabilization [23], and modeled treatment probability (propensity score) with logistic regression. To avoid large variance in the resulting estimands, we used weight trimming for percentile range 1%-99% [24,25].

#### Testing for Imbalance

We tested the imbalance between two, possibly weighted, cohorts (Step 4) by computing the absolute standardized difference [23] for each identified major confounder:

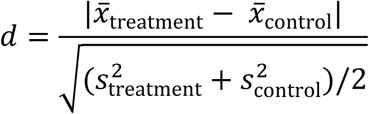

where 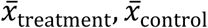 are the feature means in the two treatment groups, and 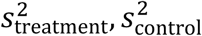 are the corresponding sample variances. We referred to the cohorts as *balanced* if for all major confounders *d* ≤ 0.2 [26].

#### Increasing Positivity Likelihood

Emulating RCTs requires satisfying the positivity condition: each patient in the trial has a positive probability of receiving either the trial drug or the alternative treatment. A failure to find balancing weights (Step 5) may indicate a violation of this condition. To increase the likelihood that the positivity condition is satisfied (Step 6), we excluded patients whose propensity scores lay outside the overlap of the treatment and control cohorts [27].

#### Effect Estimation

The Causal Inference Library provides two different methods for estimating treatment effects (Step 11), based on: (i) balancing weights, and (ii) outcome prediction. The former method estimates the expected outcome for each treatment, *a* ∈ {trial drug, alternative treatment}, using data reweighting. For event-based outcomes, we use a Kaplan-Meier estimator that reweights patients at each time unit to adjust for both confounding and selection biases:

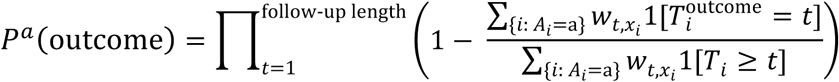

where *i* denotes a patient, *A*_*i*_ denotes the assigned treatment, 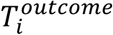 denotes the time of first outcome event, *T*_*i*_ denotes the minimum of censoring (i.e., end-of-treatment compliance) and outcome event times, and 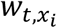 is the computed balancing weight in time *t*. In the PD case study, the time unit was one month (30 days).

The second method predicts the expected outcome for a treatment for each patient in the trial and then estimates the overall expected outcome for the treatment by taking the average:

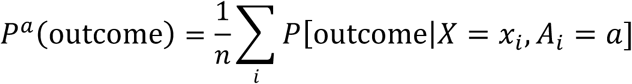

We provide complete details of these methods in the Supplementary Material. In the PD case study, we applied both methods for computing effects and analyzed their agreement.

#### P-values Estimation

We evaluated the P-value of an effect (Step 12) using a bootstrap estimate of its standard error and assuming the distribution of a sample effect is close to normal [22]. To account for multiple testing, we controlled for the false discovery rate (FDR) using the method of Benjamini and Hochberg [28]. Adjusted P-values ≤ 0.05 were considered statistically significant.

## Results

We identified ∼106,000 patients in MarketScan and ∼89,000 patients in Explorys as eligible for our emulated PD trials. To get a notion of the differences in the patient population participating in the emulated trials in each database, we compared the random-drug control cohorts in these databases. This comparison revealed many similarities, such as the average age (∼75.5 years), percentage of women (43%-45%), and the fraction of patients with public insurance (82%-85%). In both databases, dementia was the most prevalent outcome during the two-year follow-up (37-45%), followed by fall and psychosis (17-26% and 10-15%, respectively). There are also notable dissimilarities between the two databases; the most prominent is the average total patient time in database, which was more than twice as long in Explorys compared to MarketScan.

Table 1 provides statistics on the trials and results in the Explorys and MarketScan databases. Overall, we tested the effect of 259 drugs on psychosis, dementia, and fall in 1453 emulated trials, using different controls and databases. There are fewer trials with an ATC-L2 setting due to smaller control cohorts or missing ATC. For most (82%-94%) of the drugs, the RCT emulator successfully generated balancing weights in both databases (since confounders are selected based on their association with the outcome, balancing rates vary between outcomes). Despite the greater statistical power of random-drug control cohorts, we obtained more significant results using ATC-L2 control cohorts. Only 4 (0.3%) of the 1,453 trials ended with significant beneficial effects at FDR 5% by the two causal estimation methods and in both databases. Of these 4 trials, 1 drug is currently indicated for PD and the remaining 3 are repurposing candidates for treating the disease or its symptoms.

**Table 1.** Summary of trials in Explorys and MarketScan for the PD case study. Emulated trials correspond to drugs with balanced treatment and control cohorts in both Explorys and MarketScan (percentage out of the tested drugs is shown in parentheses). An identified drug’s estimated effect reduces the prevalence of the corresponding PD outcome at FDR <0.05 in both Explorys and MarketScan.

| Outcome | Control cohort | Explorys and MarketScan trials and results |  |  |
| --- | --- | --- | --- | --- |
|  |  | Tested drugs | Emulated trials (%) | Identified drugs |
| Dementia | random drug | 223 | 183 (82%) | 0 |
| Dementia | ATC-L2 | 218 | 205 (94%) | 2 |
| Fall | random-drug | 259 | 219 (85%) | 0 |
| Fall | ATC-L2 | 247 | 228 (92%) | 1 |
| Psychosis | random-drug | 259 | 218 (84%) | 0 |
| Psychosis | ATC-L2 | 247 | 214 (87%) | 1 |
| Total | random drug | 247 | 247 (100%) | 0 |
| Total | ATC-L2 | 259 | 259 (100%) | 4 |

We next applied a meta-analysis of estimated effects and assessed the level of agreement between the two different causal inference methods, namely balancing weights and outcome models. We observed (Figure 3) strong and significant correlations between the effects estimated by the two causal inference methods (focusing on drugs where at least one of the estimated effects is significant at FDR of 5%). These correlations appear to be stronger in Explorys than in MarketScan.

**Figure 3.**
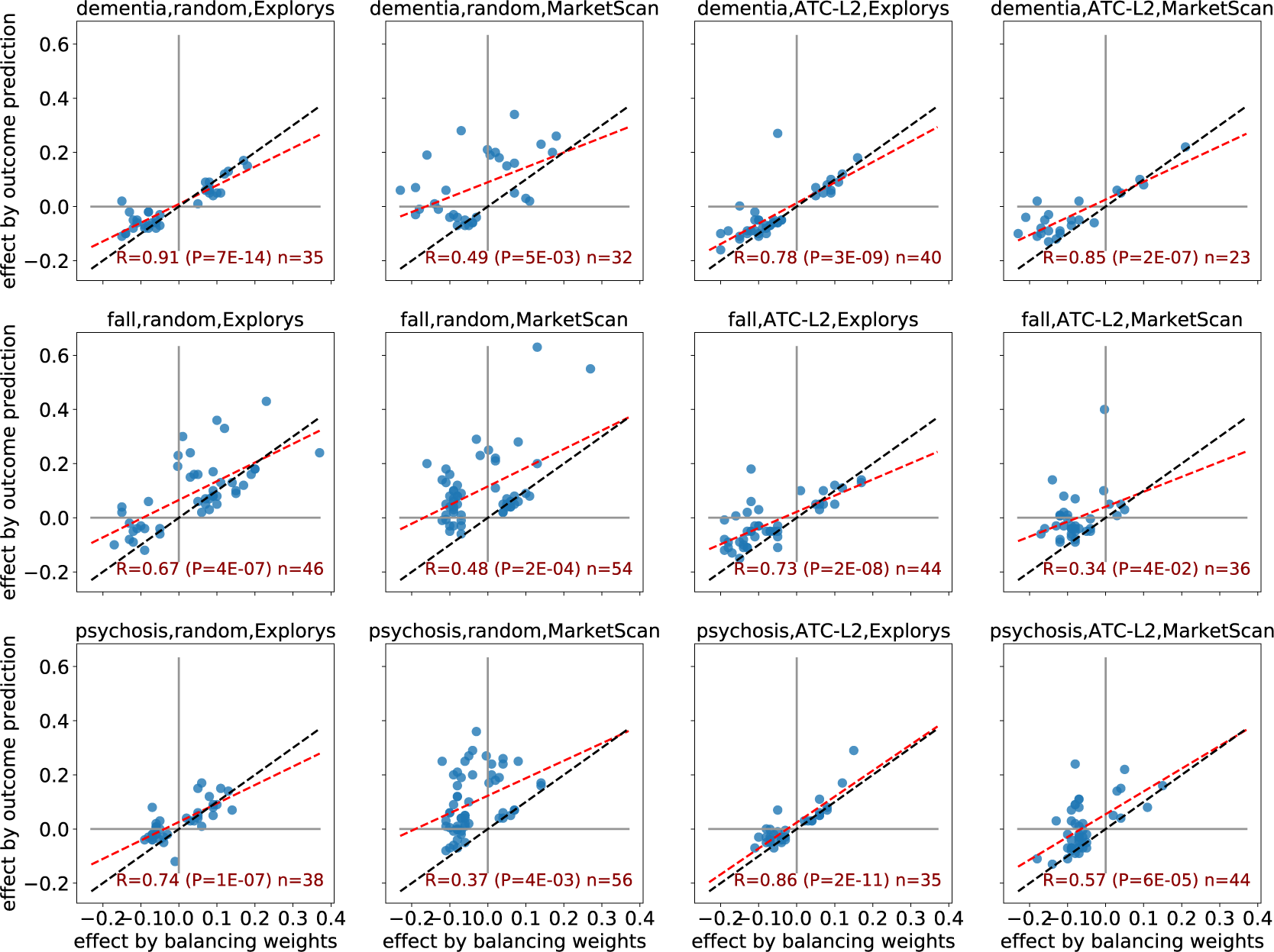
A comparison of estimated effects: balancing weights vs. outcome prediction. Each chart shows a different setting of the trial with respect to the outcome (dementia, fall, and psychosis; rows), control cohort (random and ATC-L2; left and right columns), and database (Explorys and MarketScan; alternate columns). Each point corresponds to a drug whose estimated effect was significant at FDR 5% by at least one of the two compared methods. The red line is the fitted least squares regression line; blue line indicates y=x.

Finally, to test the agreement between Explorys and MarketScan, we restricted the comparison to drugs that were shown to have a significant effect in both databases (Figure 4). The agreement between the estimated effects is remarkable, with a perfect match for effect sign (i.e., beneficial vs. harmful), and near equivalence in the magnitude of the effects. A comparison of the corresponding uncorrected effects shows similar, though somewhat weaker, agreement between the two databases (see Supplementary Figure 1).

**Figure 4.**
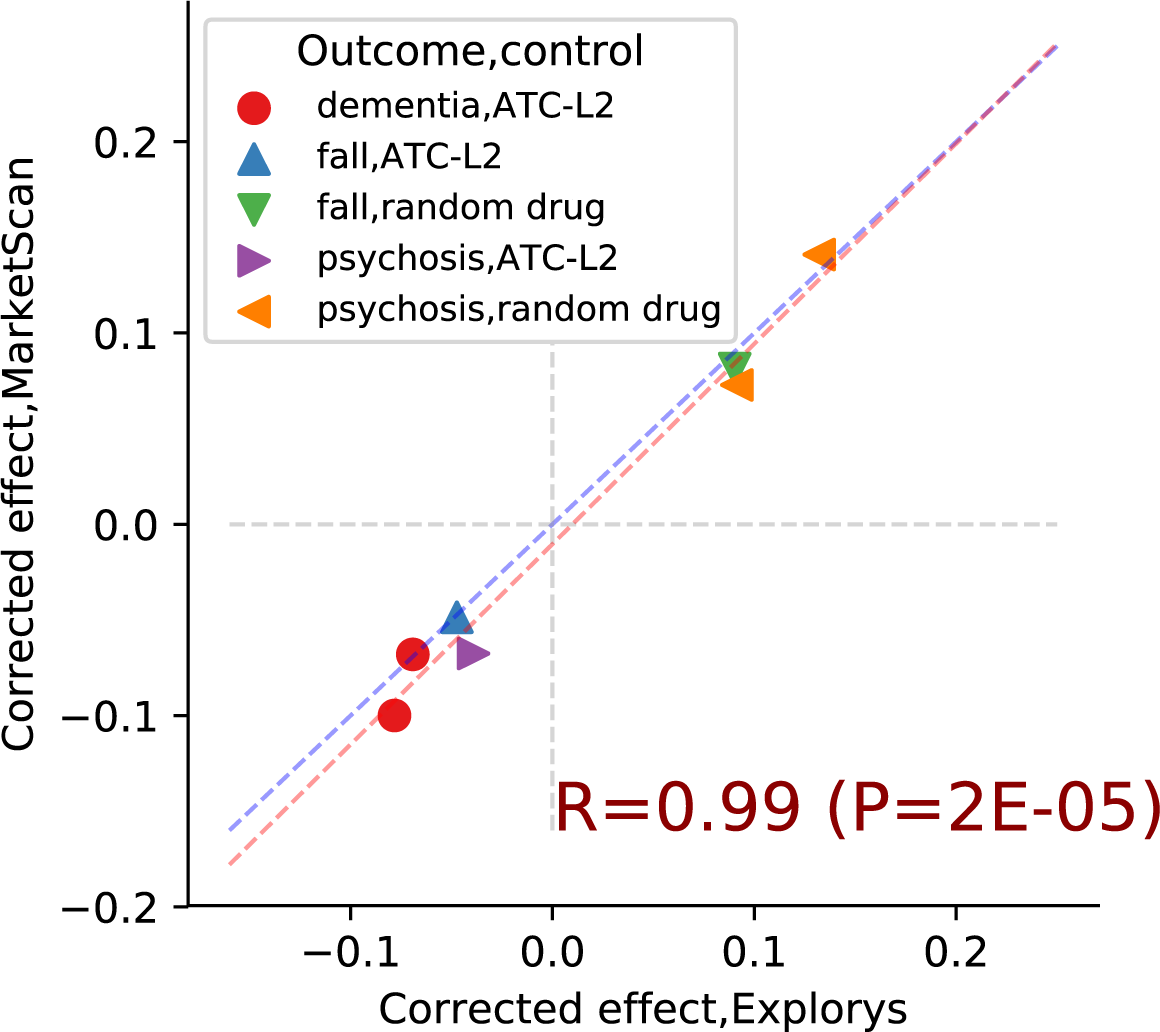
Estimated causal effects: Explorys vs MarketScan. Each point corresponds to a drug estimated to have a significant effect in both databases. Marker type represents the combination of trial outcome and control cohort; points in the first and third quadrant indicate harmful and beneficial effects, respectively. Red line is the least squares regression line, blue line is y=x.

## Discussion

A correct inference of causal effects must involve a subject-matter expert, who is also aware of the data-generation process [29,30]. Our framework allows easy injection of domain knowledge into the implementation of the emulated trials; specifically, the formulation of outcomes and hypothesized confounders [20]. There are potentially many sources of bias in healthcare data [31], leading to a large set of hypothesized confounders. However, adjusting for a high-dimensional confounder set may result in non-positivity, high-variance estimates of the effect, and over-adjustment bias [21,32]. Our framework takes a combined approach by extracting a very large number of features suspected as confounders and then applying a confounder selection step (Procedure 1, line 1). We applied a strategy that selects potential confounders based on their statistical association with the outcome [21], but other methods for confounder selection, for example [33,34], may be considered as well. While most previous drug repurposing studies that utilized observational data tested specific drug repurposing hypotheses [11,35–37], our framework screened hundreds of potential candidates. It is unique in the amount of considered confounders and the flexibility of their definition, as well as in accounting for actual treatment duration and correcting the selection bias caused by factors affecting both treatment duration and the monitored outcomes.

Another novel aspect of our framework is the random-drug control setting, where the patients in the treatment cohort are compared to other patients who start any treatment. Under this setting it may be easier to compare the estimated effects of different drugs, as all drugs are tested against a similar set of alternative drugs, representing the “background” drugs prescribed to the population under study. Furthermore, these control cohorts are relatively large, potentially increasing the statistical power of the trials. The ATC-based control setting allows the user to choose the ATC level that defines the alternative treatment. The higher the ATC level, the more closely related the pharmacological and chemical properties of the trial drugs and their alternatives, potentially ensuring a greater resemblance, or match, between the patients in the treatment and control cohorts [38]. On the other hand, as the estimated effect is a comparative measure, it may be obscured when the trial and alternative drugs similarly affect the measured outcome.

The described framework is customizable and extendible, as its components can be configured to use alternative implementations to the ones described above. A central modifiable component is the causal effect estimation method. In the PD case study, we tested two distinct approaches for causal effect estimation: balancing weights and outcome prediction. A straightforward extension is to use doubly robust methods [39], which combine the two previous approaches. Furthermore, we obtained balancing weights using the classical inverse probability weighting (IPW) method, which suffers from large estimation variance and is sensitive to model misspecification. There are many alternative methods for IPW [40–46], and each of these methods can be plugged into our framework. Using different inference methods allows the reader to evaluate the sensitivity of identified effects to modeling decisions, as suggested by [31]. Alternative implementations to other algorithmic steps, for example confounder selection, may provide an even more comprehensive evaluation of the obtained results and their robustness.

Extending the definition of a tested drug beyond single ingredients could increase the power to discover new drug candidates. In the current study, we tested only individual active ingredients, corresponding to specific molecules. Focusing on individual drugs may overlook significant effects shared by multiple similar molecules whose independent analysis lacks statistical power. To overcome this issue, we can define the set of tested drugs to be families of related molecules (e.g., using the ATC drug classification system). Similarly, we may consider drug combinations to obtain insights on synergetic effects of molecules.

We note several possible directions for extending our framework. Currently, we estimate the average effect for the entire population, although effects may be heterogenous with large differences across population strata. Identifying the sub-populations that benefit the most from each given drug (see [47] for potential approaches) could focus drug development efforts. Other directions for future work include supporting time-varying confounders and treatments to better capture temporal causal trends, incorporating drug dosage in the analysis, and inspecting the effect of inactive drug ingredients.

## Conclusion

We presented a flexible framework for high-throughput identification of drug repurposing candidates that efficiently emulates hundreds of RCTs from observational medical data to estimate the effect of on-market drugs on various disease outcomes. Naturally, the generated hypotheses require clinical analysis and experimental validation, but the significant agreement across databases and methodological approaches is encouraging. Notably, our framework may augment other *in silico* approaches [4] that leverage drug- or disease-related characteristics to identify promising drug repurposing candidates.

## Data Availability

The data analyzed in this paper is available in (i) IBM Explorys Therapeutic Dataset and (ii) IBM MarketScan Research Databases. These data does not include information on clinical trials.

## Footnotes

1 Alternative ways to measure the effect are the ratio and odds-ratio of *P*^trial drug^(outcome) and

## Notes

### Competing Interest Statement

The authors have declared no competing interest.

### Funding Statement

Study was done in IBM Research.

